# Recommendations for implementing the INTERACT3 care bundle for intracerebral hemorrhage in Latin America: a Delphi Study

**DOI:** 10.1101/2024.03.06.24303893

**Authors:** Ma.Ignacia Allende, Paula Muñoz-Venturelli, Francisca Gonzalez, Francisca Bascur, Craig S. Anderson, Menglu Ouyang, Báltica Cabieses, Alexandra Obach, Vanessa Cano-Nigenda, Antonio Arauz, LATAM INTERACT3 Consensus Statement Panel

## Abstract

**Background and aims:** The third Intensive Care Bundle with Blood Pressure Reduction in Acute Cerebral Hemorrhage Trial (INTERACT3) showed that the implementation of a care bundle improves outcomes after acute intracerebral hemorrhage (ICH). We aimed to establish consensus-based recommendations for the broader integration of the care bundle across Latin American countries (LAC).

**Methods:** A 3-phase Delphi study allowed a panel of 32 healthcare workers from 14 LAC to sequentially rank various statements that commenced with 43 statements relevant to 7 domains (training, resources and infrastructure, education of patients, blood pressure, temperature, glycemic control, and reversal of anticoagulation). The pre-defined consensus threshold was 75%.

**Results:** A total of 55 statements reached consensus by the third round, which included 12 new statements that emerged through rounds. The highest-ranked statements in each domain emphasized critical aspects with successful implementation requiring a minimum level of resources to be made available. Key priorities included the continuous training of all healthcare workers involved in the management of ICH, establishing protocols aligned with available resources, and a collaborative inter-disciplinary approach to care being supported by institutional networks. Statements related to anticoagulation reversal were given the highest priority, which highlighted interest in the topic but limited availability of treatment in the region.

**Conclusions:** Consensus statements are provided to facilitate integration of the INTERACT3 care bundle to reduce disparities in ICH outcomes in LAC.

## Introduction

Intracerebral hemorrhage (ICH) is not as common as ischemic stroke but makes a disproportionately higher contribution to the loss of productive life from stroke due to its high rates of death and disability, worldwide.^1,2^ The third INTEnsive Care Bundle with Blood Pressure Reduction in Acute Cerebral Hemorrhage Trial (INTERACT3) was an international, multicenter, stepped-wedge, cluster-randomized, controlled clinical trial that showed for the first time, that active care through a care bundle including time- and target-based protocols involving early intensive blood pressure (BP) lowering (achieving systolic BP <140 mmHg), glycemic control (achieving 6.1-7.8 mmol/L and 7.8-10.0 mmol/L without and with diabetes mellitus, respectively), treatment of pyrexia (achieving temperature level <37.5 °C), and reversal of anticoagulation (achieving an international normalized ratio [INR) <1.5), within 1 hour of the initiation of treatment and to be maintained for 7 days, led to significantly better functional recovery, lower mortality, and improved quality of life at 6-months.^3^ However, challenges exist in the broader implementation of this care bundle in ICH.

Latin American countries (LAC) had the fourth largest burden of stroke in 2019, with ICH being a major contributor due to the high prevalence of hypertension, obesity, and various barriers to integrate evidence-based care.^1,4^ LAC poses unique challenges in implementing new treatment strategies, as the healthcare systems are deeply fragmented, often with little coordination between discipline groups.^5^ Inequitable access to healthcare that is coupled with substantial budget constraints, impedes the capacity to address healthcare demands arising from social and demographic shifts. Structural disparities between social groups, limited availability of universal healthcare coverage, and diversity in culture and health literacy in the region present considerable challenges to addressing the burden of stroke as well as other common conditions.^5,6^

A better understanding of facilitators and barriers can facilitate the integration of new interventions and improve knowledge-to-practice gaps. The INTERACT3 study captured some barriers to implementing the care bundle through an embedded process evaluation. We wished to consolidate such information by generating structured strategies specific to LAC. Herein, we report our efforts to establish recommendations to the implementation of the INTERACT3 care bundle in hospitals across LAC.

## Methods

### Study design

We performed a 3-phase online Delphi process with the aim of obtaining expert recommendations for implementing the INTERACT3 care bundle in LAC. The goal of the Delphi methodology is to achieve expert consensus by employing semi-structured questionnaires with open-ended questions and controlled assessment and feedback to pre-defined statements. Mixed methods-quantitative and qualitative-analysis are undertaken until an accord is established and summarized.^7,8^ The method is widely used to establish recommendations for patient care and public health, such as during the COVID-19 pandemic.^9–11^ Our study conformed to the Recommendations for the Conducting and Reporting of Delphi Studies (CREDES) (Table S1).^12^

### Generation of the Delphi survey

Two of us (MIA and FGM) conducted a comprehensive review of the INTERACT3 process evaluation that was limited to LAC (results are unpublished) to formulate 43 statements to include in the first round. These statements were thoughtfully categorized into seven domains: (i) training and education (6 statements); (ii) human resources and infrastructure (5 statements); (iii) early intensive BP control (7 statements); (iv) strict glucose control (12 statements); (v) body temperature control (6 statements); (vi) rapid reversal of anticoagulation (3 statements); and (vii) education for patients and family members (4 statements). Two survey versions were designed in English and Spanish to accommodate linguistic diversity. Extensive discussions were undertaken of the domains and individual statements over two meetings. The rounds underwent careful examination for clarity, coherence, and linguistic refinement in neutral Spanish (VCN) and English (MO and CA) to produce the final questionnaire.

### Selection of panelist

We used an iterative sampling approach to identify panelists (Figure 1). Potential participants were identified from known specialist contacts of the research group and through snowball recruitment. Invitations to participate were extended through an online recruitment letter that outlined the study objectives. Panelists were offered compensation for their time (USD$100) upon completion of the study.

**Figure 1.**
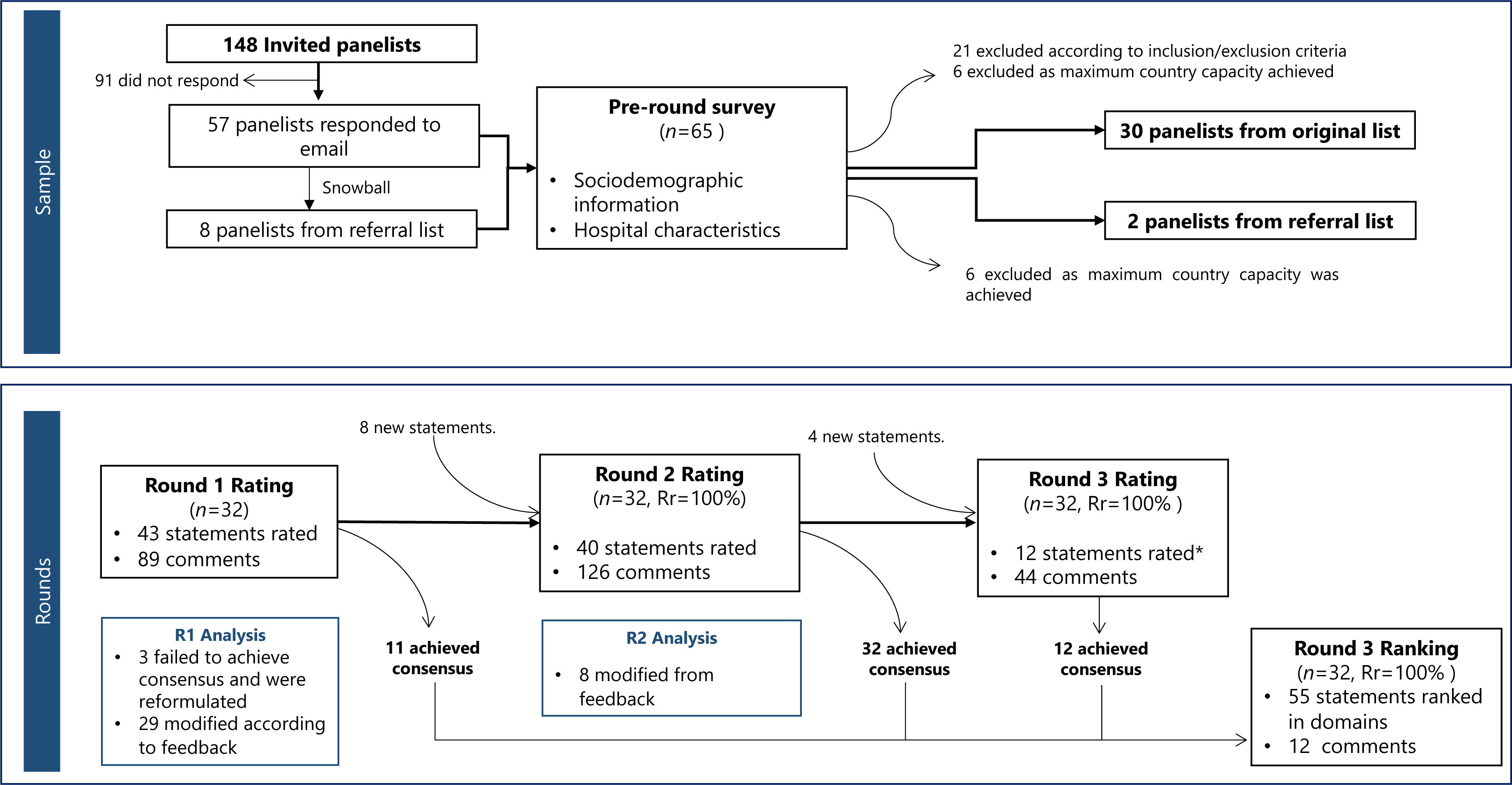
Flowchart of sampling and Delphi rounds. Iterative sampling approach resulted in 32 panelists in all rounds. Rr = Response rate. *R3 analysis not shown since all 12 statements achieved consensus.

To assess the expertise of invited participants, a survey collected demographic and hospital characteristic data in the initial recruitment letter. Eligibility for participation included the following criteria: being a healthcare professional with a minimum of 5 years of experience in a respective field; having at least one year of experience in a public health facility; and being currently involved in the care of ICH patients, including the hyperacute care phase of stroke. People working in a facility without computerized tomography or an emergency area were excluded.

We defined the required sample size as 32 participants to align with similar studies where 30-40 was considered adequate to define common themes.^13–15^ Participants were selected to a maximum of 4 panelists from each of 16 LAC for representation.

### Data collection

We pre-specified 3 rounds to ensure consensus and mitigate drop-out, based on previous research indicating results diminish or stabilize at this stage.^16^ A REDCap® platform was used to generate and distribute surveys.^17,18^

Each round lasted 14 days, and an average of 3 reminders was needed to prompt survey completion. Between rounds, the research group allotted 3 weeks to analyze responses and create the subsequent round; this allowed a separation of 5 weeks between rounds. In the first round (R1), panelists were asked to rate each of the statements according to their level of agreement on a descending 4-point Likert scale (strongly agree [SA], agree [A], disagree [D], and strongly disagree [SD]). Following each statement and dimension, panelists could provide comments and suggest edits for the current statements or propose additional ones.

The pre-defined consensus threshold was set at 75% agreement (SA or A) for retention of a statement, as defined elsewhere.^9,15,19^ Statements below this threshold were modified according to feedback from panelists, and some statements were modified if they failed to reach 75% of SA from panelist feedback.

In the second round (R2), statements with modifications and any new ones suggested by panelists, were included in the survey. Statements that achieved consensus agreement were removed and only include in the final ranking section in round 3 (R3). Panelists were provided with percentage agreement in R1 and relevant unidentified citations. As in R1, they were provided with open comment boxes at the end of each statement and dimension.

R3 followed a similar dynamic process, but this time the panelists had to rank the importance of all retained statements on a 9-point Likert scale, from 1 ‘most important’ to 9 ‘least important’. Ultimately, they could indicate which statement they considered should be removed from the final report; this applied to all statements that received a 75% agreement for exclusion. The goal was to ensure that the final report only included statements with a substantial level of importance. Anonymity was maintained throughout the Delphi rounds.

### Data analysis

Quantitative data were analyzed using descriptive statistics. Statements results were summarized using percentage of agreement, and consensus was achieved if ≥75% of combined ‘SA’ and ‘A’ responses were obtained. For the ranking section, median and percentage of statements rated ≥3 were reported. STATA version 18 was used in all statistical analyzes.

Qualitative data were analyzed (MIA and FGM) using a simple content analysis from the comments gathered in each round. Group consensus was used to decide on the requirement to modify existing statements or consider new statements that emerged in each round. Verbatim quotes from participants are presented to support the findings.

### Ethical approval

The study was approved by the Scientific Ethics Committee of Clínica Alemana-Universidad del Desarrollo (number 2023-68). All panelists provide an online informed consent before participation in the study.

## Results

The panelist group comprised 32 participants (mean age 37.1±5.8 years) from 14 LAC (Figure 2) who were predominantly neurologists (87.5%) with a mean of 9.7±3.7 years of experience. Table 1 provides details of their other characteristics. They responded to all 3 rounds (Figure 1). In R1, of the 43 statements, 3 failed to reach the pre-defined agreement of 75%, 29 were modified based on weak agreement, and 8 new statements emerged from the panelists. Thus, the 11 statements that achieved consensus were not modified nor passed on to R2.

**Figure 2.**
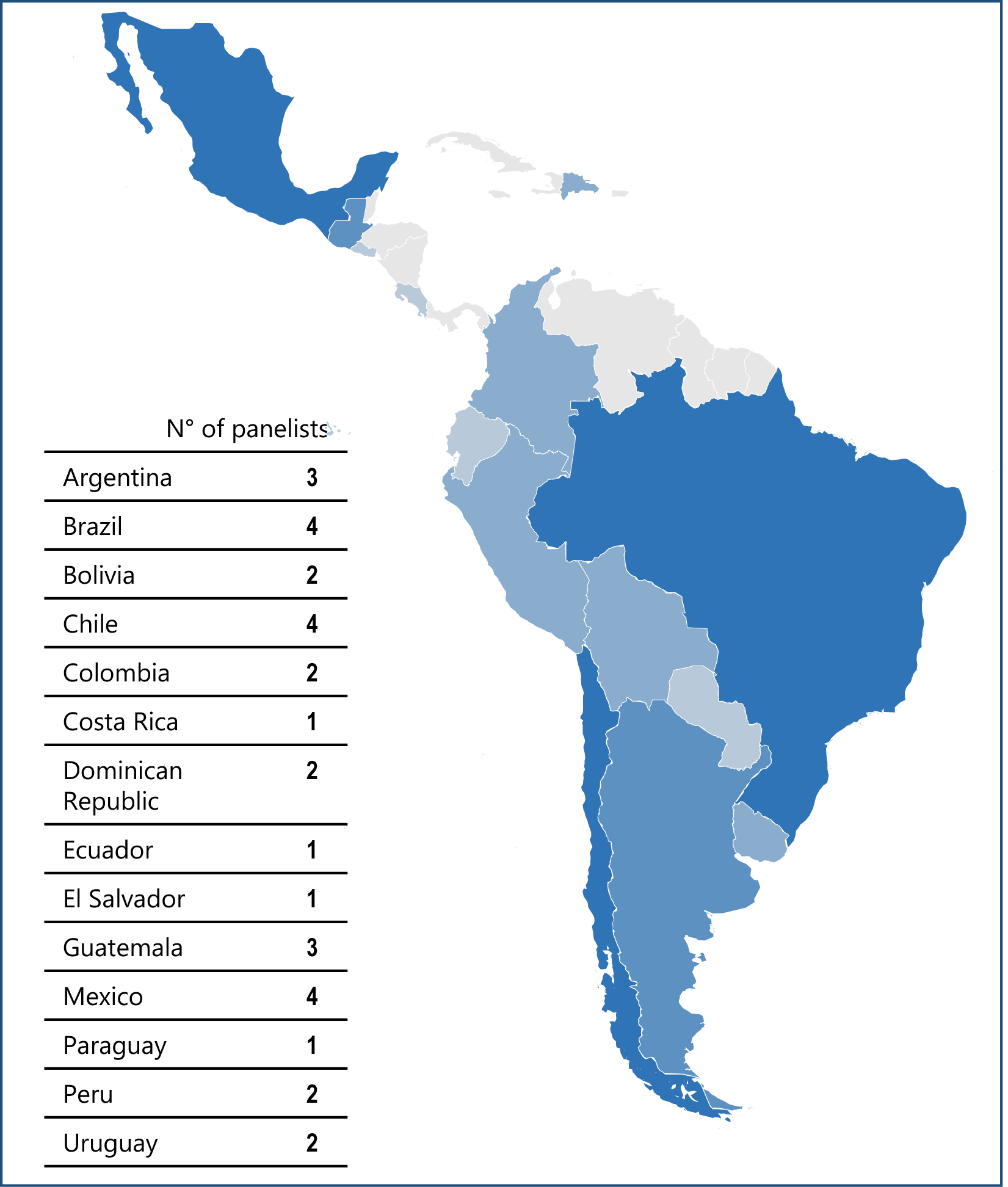
Distribution of panelists in Latin America.

**Table 1.** Baseline panelist characteristics.

|  | Participants<br>(n = 32) |
| --- | --- |
| *Sociodemographic* |  |
| Male | 21 (65.62) |
| Age, yr | 37.1 ±5.8 |
| Income of country(*) |  |
| Low-middle income | 2 (6.25) |
| High-middle income | 24 (75) |
| High income | 6 (18.75) |
| Profession |  |
| Medical Doctor | 31 (96.88) |
| General practitioner | 1 (3.13) |
| Specialist |  |
| Neurology | 28 (87.50) |
| Internal Medicine | 1 (3.13) |
| Urgenciology | 1 (3.13) |
| Physiotherapist | 1 (3.13) |
| Experience |  |
| Years in profession, mean ±SD | 9.71 ±3.70 |
| Years in ICH patient care, mean ±SD | 7.62 ±2.76 |
| Years in Public Health System, mean ±SD | 8.12 ±4.05 |
| INTERACT3 investigator | 6 (18.75) |
| *Panelist hospital* |  |
| Location |  |
| Urban | 30 (93.75) |
| Semi-urban | 2 (6.25) |
| Rural | 0 (0) |
| Health Sector |  |
| Private | 6 (18.75) |
| Public | 25 (78.13) |
| Other | 1 (2.70) |
| Computerized tomography available | 32 (100) |
| Service Units |  |
| Emergency department | 32 (100) |
| Neurosurgery | 27 (84.38) |
| Intensive Care Unit | 32 (100) |
| Stroke Unit | 16 (50) |
| Laboratory facilities | 31 (96.88) |
| Stroke Protocol | 26 (81.25) |
| Inclusion of ICH treatment | 17 (65.38) |

Data are n (%) or mean±SD
(\*)Income of countries based on the World Bank classification system: high-income includes Chile and Brazil; high-middle-income includes Argentina, Brazil, Colombia, Costa Rica, Dominican Republic, Ecuador, El Salvador, Guatemala, Mexico, Paraguay, and Peru; low-middle-income include Bolivia.
Abbreviations: ICH= intracerebral hemorrhage. INTERACT3=The third INTensive Care Bundle with Blood Pressure Reduction in Acute Cerebral Hemorrhage Trial.

In R2, of the 40 statements that achieved consensus, 8 were modified due to lack of SA consensus and 4 new statements were created. The 32 statements that achieved consensus were not modified and did not pass to R3. Thus, the final round involved the rating of 12 statements and by the end, all statements achieved consensus. The panelists also had the opportunity to rank all 55 statements considered throughout the rounds within each domain. As there were no statements with 75% consensus agreement for exclusion from the final report, the following section presents all the statements in the final round, with quotes to emphasize relevance.

### Training and education

Statements concerning national initiatives were the highest ranked and achieved substantial consensus. For example, the need to create a ‘stroke code’ protocol for the early treatment of ICH patients (SA 93.8%; ranking 1.34±1.45) (Table S2). Moreover, education to the general public was to emphasize the importance of symptom recognition and need to promptly call emergency services (SA 84.4%; ranking 1.46±1.31).

> “One of the main limitations in providing and applying the bundle of care is that symptoms are not recognized in time, and as a result, individuals do not seek medical attention.”

Emphasis was made to provide training to all healthcare workers involved in the management of ICH patients (SA 93.8%; ranking 1.34±1.45), without prioritizing certain groups over others.

> “Although the background of the neurology team is usually better, I believe that training should be given to all staff and not prioritizing non-neurologist staff”

The high rotational nature of staffing in emergency rooms was highlighted as a barrier to effective implementation. Thus, efforts should be made towards continuous education of staff.

### Human resources and infrastructure

The key concern was the provision of training to healthcare workers (SA 90.6%; ranking 1.09±0.29) (Table S3), and the necessity for cohesion between different discipline units.

> “All personnel who treat patients with intracerebral hemorrhage should be on the same page.”

Assurance over the allocation of minimum resources in countries (SA 81.3%; ranking 1.43±0.84) was also deemed a high priority. Importance was also given to statements that highlighted the need for a collaborative approach through the creation of communication pathways between units to ensure prompt notification of imaging results (SA 68.8%; ranking 1.71 ±1.05); and continuity of treatment for patients transferred to other units (SA 68.8%; ranking 1.75 ±1.27).

> “In-hospital delays due to miscommunication are higher than we generally assume.”

### Early intensive BP control

Adherence to the strict monitoring protocol used in INTERACT3 was given the highest priority (Table S4). Despite a good overall correlation between rating and ranking being achieved, this statement had the highest combined disagreement in the domain (Combined Disagreement [CD] 15.7%, ranking 1.28±0.58). Although the panelists agreed on the minimum set of BP measurement used in the INTERACT3 protocol, monitoring requirements should be based on individual requirements.

> “Monitoring may need to be invasive and continuous or stricter in the first 72 hours in patients and even more so if they are being managed with intravenous treatment.”

> “It should be understood that this statement speaks of as a minimum so it is clear that it leaves the possibility of individualizing monitoring according to the situation and severity of each patient.”

Ensuring that there is a minimum level of resources being readily available, such as intravenous antihypertensive drugs, was ranked highly (SA 81.3%; ranking 1.31±0.64). Some panelists said this was the main barrier to implementation. Given challenges in the allocation of healthcare resources, initiatives to provide treatment protocols aligned with local resources was encouraged (SA 87.5%; ranking 1.37±0.55). Emphasis was also placed on providing evidence-based protocols that are periodically updated.

### Strict glucose control

Recommendations in this domain were driven by concerns over the requirement for insulin pumps in the INTERACT3 protocol (Table S5). Thus, the highest ranked recommendation was the necessity to include practical training sessions on glycemic management, despite having a relative low agreement (SA 59.4%; ranking 1.5±0.80). Moreover, panelists expressed concern over the widespread unavailability of insulin pumps in LAC; these are generally reserved for patients with extremely poor glucose control. Panelists agreed that specific protocols (SA 87.5%, ranking 1.68±0.89) and communication channels with endocrinologists and pharmacists (SA 40.6%, ranking 1.75±0.76) should be established for complex patients.

> “In complicated scenarios, there should be guidelines to help in the optimal monitoring and control of glycemia”.

Another highly ranked recommendation was the need for assurance over the strict measurement of blood glucose level (BGL) (SA 71.9%; ranking 1.59±0.87). Compared to the BP monitoring domain, there was less agreement over the glycemic protocol used in INTERACT3 (SA 31.3%; ranking 3±2.01). This primarily stemmed from its use in the stable non-diabetic patient, where consideration was recommended towards using a more relaxed monitoring of BGL as resources are limited.

> “If the patient is not diabetic or does not have elevated blood sugar at the onset, I do not think it is advisable to spend resources”.

### Body temperature control

Emphasis was on training of healthcare workers (SA 75%; ranking 1.53±1.13) (Table S6). Monitoring of patients was also considered necessary (SA 59.4%; ranking 1.75±0.84).

> “Focus on proper monitoring, and temperature control. This is a point that is often overlooked.”

### Rapid reversal of anticoagulation

Key aspects were the availability and affordability of modern treatments in this increasing important topic (Table S7). The utmost priority was assurance over the availability of anticoagulation reversal agents (SA 81.3%; ranking 1.21±0.49).

> “Establish protocols where there is no need to have the approval of different sectors to allow for the administration of reversing agents.”

Another aspect was the need for strategies to ensure 24-hour availability of INR measurement as part of a collaborative approach, supported by networks between healthcare facilities (SA 81.3%; ranking 1.21±0.49). The need to provide training was also highlighted (SA 87.5%; ranking 1.31±0.50).

> “There is fear with patients who experience bleeding on anticoagulants. Doctors suspend treatment, and there is a fear of restarting due to recurrence. Workshops on this topic are important because not everyone is familiar with it.”

### Education for patients and family members

Use of plain language to educate patients and their families (and/or caregivers) was considered a priority (SA 90.6%; ranking 1.37±0.75) (Table S7). Given that need for frequent monitoring of the care bundle, panelists emphasized the need to educate relatives on the benefits of this approach for ICH patients (SA 78.1%; ranking 1.62± 0.75), whilst also avoiding projecting unrealistic expectations.

> “It should be clear, without providing false expectations, but without discouraging, as this can limit the opportunity for improvement.”

### Overall priorities

The statement that received the highest ranking across all domains was the provision of training for all healthcare workers, which emphasized the necessity for teamwork during implementation of the care bundle (SA 90.6%; ranking 1.09± 0.29). Anticoagulation reversal emerged as the domain with the highest-prioritized statements, followed by BP control (Table S4 and S5). Within these domains, the statements that received the highest rankings were those related to resource allocation and education.

## Discussion

Our study provides fresh perspectives on areas that need to be addressed to ensure successful integration of the INTERACT3 care bundle across hospitals in LAC. High-priority domains were BP control, anticoagulation reversal, and resourcing, with a focus on education and training. Blood glucose and temperature control domains had lower rankings and received less consensus overall. Use of the Delphi technique allowed us to identify recommendations from a comprehensive perspective of different clinician stakeholders in LAC.

A common perspective was the need to ensure that a minimum level of resources would be made available to achieve effective integration of the care bundle. Concerns were raised over the shortage of antihypertensive drugs and reversal anticoagulation agents, which panelists deemed as a critical barrier in being able to achieve effective implementation of key components of the care bundle. This is consistent with previous studies in which contextual factors, such as financial resource constraints, are among the most frequently mentioned barriers.^20^ Other recommendations related to resources, including the availability of glucometers, thermometers, and BP monitors, did not receive as high a level of ranking, possibly because they reflected a lower priority of need. Recognizing the substantial challenges in the allocation of healthcare resources in LAC, the panelists acknowledged the importance of developing simple and efficient protocols. However, prioritizing the supply of core items, such as the items ranked higher, was deemed a crucial point in resource-constrained settings in LAC.

In common with other country-level recommendations, the generation of a ‘stroke code’ emerged as another critical aspect. While there has been a global focus on prompt implementation of reperfusion therapy for acute ischemic stroke over recent decades, no such urgency has been uniformly applied to patients with ICH, in part due to the lack of any proven treatments and also due to the natural history of a poor outcome. Indeed, while 90% of the panelists reported having a “stroke code” protocol in their workplaces, only two-thirds had an integrated ICH management pathway. The panelists emphasized a pressing need to address this gap given that robust evidence is now emerging that time makes a difference to prognosis in ICH. The non-aggressive approach of clinicians to ICH is further reflected in the common use of low threshold to withdraw active care in these patients.^21^ Training on the benefits of the INTERACT3 care bundle was considered a means of addressing premature use of do-not-resuscitate orders and clinician-misjudgment over prognostication.^22^ Hence, inclusion of an ICH pathway and the training of all healthcare workers on the INTERACT3 care bundle were considered pivotal first steps in reshaping the perspectives of healthcare professionals on ICH management.

It is important to recognize that since ICH is the most severe form of stroke, many patients are unable to actively participate in the decision-making process.^23^ In such overwhelming scenarios, misinterpretation of prognosis by surrogates decision-makers, most often being immediate family members, can lead to misunderstandings, emotional distress and conflict, in response to early decisions to forego treatment.^23,24^ In order to provide goal-concordant care, the panelists emphasized the vital role that counselling of relatives on prognosis has on ensuring a clear message is received through the use of simple language without instilling false hope.

Our findings further highlight the need for a collaborative approach in stroke care by the creation of communication channels between professionals and the creation of networks between healthcare institutions. The dimensions for a collaborative approach were for strict glucose control and anticoagulation reversal, reflecting the limited availability of treatments and limited knowledge of approaches in those areas. The panelists were reluctant to promote the use of insulin pumps, which is consistent with barriers found in the use of the fever, hyperglycemia, and swallowing dysfunction protocols used in the Quality in Acute Stroke Care (QASC) study.^25^ Similarly, the process evaluation undertaken among Chinese participants in INTERACT3 found that blood glucose control was the most complex intervention to implement, and that collaboration and consultations with endocrinology were key facilitators aligns with our findings.^26^ During the Delphi rounds, use of subcutaneous insulin was often mentioned but it did not reach a high ranking, further emphasizing the need for training and protocols in the region.

Given the relative low use of warfarin anticoagulation in Latin America,^27^ and thus a lack of exposure to anticoagulation reversal, adequate supply of medications alone would unlikely lead to effective implementation of the care bundle. Continuous training on drug reconstitution, organizational support, and ensuring 24-hour availability on INR measures through collaborative networks, was contemplated as being important for successful integration. Other studies have emphasized these being key steps in reducing door-to-needle time for anticoagulation reversal,^28^ and are arguably even more relevant in the era of reversal agents for direct oral anticoagulants.

Our study has several strengths. The Delphi method was used to evaluate recommendations for implementation of the INTERACT3 care bundle to enhance ICH management. The panelists from 14 LAC led to a comprehensive perspective being gained that accounted for variations in healthcare systems and cultural contexts in the region. They provided a complete response rate throughout rounds that enhances reliability of the results. However, some limitations are worth mentioning. First, despite our efforts to construct a multidisciplinary panel, it mainly comprised specialist clinicians without any lived-experience patient, family or community engagement. Although the panelists were experienced and worked in a public system, provided them with a holistic view, they may not necessarily reflect the views of junior staff who work at the front line and are involved in the day-to-day management of patients with ICH. Finally, as this study focused on LAC, the views may not be representative of other countries or healthcare systems.

In summary, the INTERACT3 bundle of care has the potential to reduce the burden of ICH in LAC, by alleviating alleviate strain on diverse and complex health systems and improving the health and wellbeing of those affected. Using the Delphi technique, our study provides a priority list of key aspects for effective integration of the INTERACT3 care bundle in the region. Key stakeholders can use these findings to devise context-specific strategies towards effective implementation.

## Data Availability

The data that support the findings of this study are available from the corresponding author,PMV, upon reasonable request.

## Acknowledgments

The authors thank the LATAM INTERACT3 consensus panel.

## Supplemental Material

List of LATAM INTERACT3 Consensus Panel

Tables S1-S7

## References

1. Feigin VL, Stark BA, Johnson CO, Roth GA, Bisignano C, Abady GG, et al. Global, regional, and national burden of stroke and its risk factors, 1990–2019: a systematic analysis for the Global Burden of Disease Study 2019. Lancet Neurol 2021;20:795–820

2. Magid-Bernstein J, Girard R, Polster S, Srinath A, Romanos S, Awad IA, Sansing LH. Cerebral hemorrhage: pathophysiology, treatment, and future directions. Circ Re. 2022;130:1204–1229

3. Ma L, Hu X, Song L, Chen X, Ouyang M, Billot L, et al. The third Intensive Care Bundle with Blood Pressure Reduction in Acute Cerebral Haemorrhage Trial (INTERACT3): an international, stepped wedge cluster randomised controlled trial. Lancet 2023;402:27–40

4. Pacheco-Barrios K, Giannoni-Luza S, Navarro-Flores A, Rebello-Sanchez I, Parente J, Balbuena A, et al. Burden of stroke and population-attributable fractions of risk factors in Latin America and the Caribbean. JAHA 2022;11:e027044

5. Atun R, De Andrade LOM, Almeida G, Cotlear D, Dmytraczenko T, Frenz P, et al. Health-system reform and universal health coverage in Latin America. Lancet 2015;385:1230–1247

6. Ruano AL, Rodríguez D, Rossi PG, Maceira D. Understanding inequities in health and health systems in Latin America and the Caribbean: a thematic series. Int J Equity Health 2021;20:94, s12939–021-01426–1

7. Okoli C, Pawlowski SD. The Delphi method as a research tool: an example, design considerations and applications. Information Management 2004;42:15–29

8. Keeney S, Hasson F, McKenna H. Consulting the oracle: ten lessons from using the Delphi technique in nursing research. Journal of Advanced Nursing 2006;53:205–212.

9. Lazarus JV, Romero D, Kopka CJ, Karim SA, Abu-Raddad LJ, Almeida G, et al. A multinational Delphi consensus to end the COVID-19 public health threat. Nature 2022;611:332–345

10. Rahaghi FF, Balasubramanian VP, Bourge RC, Burger CD, Chakinala MM, Eggert MS, et al. Delphi consensus recommendation for optimization of pulmonary hypertension therapy focusing on switching from a phosphodiesterase 5 inhibitor to riociguat. Pulm Circ 2022;12:e12055

11. Van Pottelberghe S, Heine F, Van Dooren S, Hes F, Kupper N. Barriers and facilitators for the implementation of patient-centered care in cardiogenetics: a Delphi study among ERN GUARD-heart members. Eur J Hum Genet 2023;31:1371–1380.

12. Jünger S, Payne SA, Brine J, Radbruch L, Brearley SG. Guidance on Conducting and REporting DElphi Studies (CREDES) in palliative care: recommendations based on a methodological systematic review. Palliat Med 2017;31:684–706

13. Stein J, Rodstein BM, Levine SR, Cheung K, Sicklick A, Silver B, et al. Which road to recovery? Factors influencing postacute stroke discharge destinations: a Delphi study. Stroke 2022;53:947–955

14. Regan M, Smolar M, Burton R, Clarke Z, Sharpe C, Henn C, Marsden J. Policies and interventions to reduce harmful gambling: an international Delphi consensus and implementation rating study. Lancet Public Health 2022;7:e705–e717

15. Mason B, Boyd K, Doubal F, Barber M, Brady M, Cowey E, et al. Core Outcome measures for palliative and end-of-life research after severe stroke: mixed-method Delphi study. Stroke 2021;52:3507–3513

16. López-Gómez E. El método Delphi en la investigación actual en educación: una revisión teórica y metodológica. Educación XX1. 2017;21(1):17-40

17. Harris PA, Taylor R, Thielke R, Payne J, Gonzalez N, Conde JG. Research electronic data capture (REDCap): a metadata-driven methodology and workflow process for providing translational research informatics support. Journal of Biomedical Informatic. 2009;42:377–381

18. Harris PA, Taylor R, Minor BL, Elliott V, Fernandez M, O’Neal L, et al. The REDCap consortium: Building an international community of software platform partners. J Biomed Informatics 2019;95:103208

19. Diamond IR, Grant RC, Feldman BM, Pencharz PB, Ling SC, Moore AM, Wales PW. Defining consensus: a systematic review recommends methodologic criteria for reporting of Delphi studies. J Clin Epidemiol 2014;67:401–409

20. Baatiema L, Otim ME, Mnatzaganian G, de-Graft Aikins A, Coombes J, Somerset S. Health professionals’ views on the barriers and enablers to evidence-based practice for acute stroke care: a systematic review. Implementation Sci 2017;12:74

21. Parry-Jones AR, Paley L, Bray BD, Hoffman AM, James M, Cloud GC, et al. Care-limiting decisions in acute stroke and association with survival: analyses of UK national quality register data. Int J Strok. 2016;11:321–331

22. Bailey S. The Concept of Futility in Health Care Decision Making. Nurs Ethics 2004;11:77–83

23. Gao L, Zhao CW, Hwang DY. End-of-Life Care Decision-Making in Stroke. Front Neurol 2021;12:702833

24. Hemphill JC, White DB. Clinical Nihilism in Neuroemergencies. Emerg Med Clin N Am 2009;27:27–37

25. Dale S, Levi C, Ward J, Grimshaw JM, Jammali-Blasi A, D’Este C, et al. Barriers and enablers to implementing clinical treatment protocols for fever, hyperglycaemia, and swallowing dysfunction in the quality in acute stroke care (QASC) project: a mixed methods study. Worldviews Ev Based Nurs 2015;12:41–50

26. Ouyang M, Anderson CS, Song L, Jan S, Sun L, Cheng G, et al. Implementing a goal-directed care bundle after acute intracerebral haemorrhage: process evaluation for the Third INTEnsive Care Bundle with Blood Pressure Reduction in Acute Cerebral Haemorrhage Trial Study in China. Cerebrovasc Dis 2022;51:373–383

27. Kozieł M, Teutsch C, Bayer V, Lu S, Gurusamy VK, et al. Changes in anticoagulant prescription patterns over time for patients with atrial fibrillation around the world. J Arrhythmia 2021;37:990–1006

28. Parry-Jones AR, Järhult SJ, Kreitzer N, Morotti A, Toni D, Seiffge D, et al. Acute care bundles should be used for patients with intracerebral haemorrhage: an expert consensus statement. Eur Stroke J 2023;(0)

